# Beyond HER2 expression in breast cancer: Investigating alternative splicing profiles as a mechanism of resistance to anti-HER2 therapies

**DOI:** 10.1101/2024.11.25.24317569

**Authors:** Gabriela D. A. Guardia, Carlos H dos Anjos, Aline Rangel-Pozzo, Filipe F. dos Santos, Alexander Birbrair, Paula F. Asprino, Anamaria A. Camargo, Pedro A F Galante

## Abstract

Breast cancer is a heterogeneous disease that can be molecularly classified based on the expression of hormone receptors and the overexpression of the HER2 receptor (ERBB2). Targeted therapies for HER2-positive breast cancer, including trastuzumab, antibody drug conjugates (ADCs) and tyrosine kinase inhibitors, have significantly improved patient outcomes. However, both primary and acquired resistance to these treatments pose challenges that can limit their long-term efficacy. Addressing these obstacles is vital for enhancing therapeutic strategies and patient care. Alternative splicing, a post-transcriptional mechanism that enhances transcript diversity (isoforms) within a cell, can result in isoform-encoded proteins with varied functions, cellular localizations, or binding properties. In this study, we undertook a comprehensive characterization of the alternative splicing isoforms of HER2, assessed their expression levels in primary breast tumors and cell lines, and explored their role in resistance to anti-HER2 therapies. Our results have significantly expanded the catalog of known HER2 protein-coding isoforms from 13 to 90, revealing distinct patterns of protein domains, cellular localization, and protein structures, as well as mapping their antibody-binding sites. Additionally, by profiling expression in 561 primary breast cancer samples and analyzing mass spectrometry data for translation evidence, we discovered a complex landscape of splicing isoform expression in primary tumors, revealing novel isoforms that were previously unrecognized and are not evaluated in routine clinical practice. This extends beyond the traditional profile based solely on HER2 gene expression and translation. Finally, by assessing HER2 isoform expression in cell cultures that are either sensitive or resistant to trastuzumab and ADCs (T-DM1 or T-DXd), we found that drug-resistant tumor cells shifted their expression toward splicing isoforms that lack the antibody-binding domains. Our results substantially broaden the understanding of HER2 protein-coding isoforms, revealing distinct mechanisms of potential resistance to anti-HER2 therapies, particularly ADCs, by uncovering a new dimension of splicing isoform diversity. This expanded landscape of HER2 isoforms, marked by unique domain patterns and altered antibody-binding sites, emphasizes the crucial role of alternative splicing investigations in advancing precision-targeted cancer therapies.

## INTRODUCTION

Breast cancer remains one of the most prevalent and challenging malignancies worldwide, with an estimated 2.3 million new cases diagnosed in 2022 (F. Bray et al. 2024). The heterogeneity of breast cancer at both clinical and molecular levels has compelled its classification into distinct groups, enabling more tailored treatment approaches for improving patient outcomes (Perou et al. 2000). The molecular classification of breast cancer, primarily based on the expression of hormone receptors (estrogen and progesterone) and the human epidermal growth factor receptor 2 (HER2, also known as ERBB2), has revolutionized our understanding of this disease and guided the development of HER2 targeted therapies (Sørlie et al. 2001; Prat et al. 2017).

HER2-positive breast cancers, accounting for approximately 20% of all breast cancers, are characterized by the overexpression or amplification of the HER2 gene (Wolff et al. 2013). Located on chromosome 17q12, the HER2 transcripts typically encode a 185 kDa transmembrane tyrosine kinase receptor that belongs to the epidermal growth factor receptor (EGFR) family (Moasser 2007). Regarding functionalities, HER2 overexpression leads to the constitutive activation of downstream signaling pathways, including the PI3K/AKT and MAPK, promoting tumor cell proliferation and survival (Yarden and Sliwkowski 2001). The HER2-positive breast cancer subtype is associated with aggressive tumor behavior and poor prognosis in the absence of anti-HER2 targeted therapy (Slamon et al. 1987).

The advent of HER2-targeted therapies has significantly improved the outcomes for patients with HER2-positive breast cancer. Trastuzumab, a humanized monoclonal antibody targeting the extracellular domain of HER2, was the first targeted therapy approved for both metastatic and early-stage HER2-positive breast cancer (Slamon et al. 2001; Piccart-Gebhart et al. 2005). Since then, additional therapeutic classes have expanded the treatment landscape for HER2-positive disease, including tyrosine kinase inhibitors such as lapatinib, neratinib, and tucatinib, and, more recently, HER2-targeted antibody-drug conjugates like T-DM1 and T-DXd, which combine the targeting precision of trastuzumab with potent cytotoxic agents (Verma et al. 2012; Modi et al. 2020). Altogether, these new therapeutic options have had a profound impact on the management of both metastatic and early-stage HER2-positive breast cancer, extending disease control and significantly reducing the risk of recurrence (Swain et al. 2015; von Minckwitz et al. 2017)

Therapeutic advancements have greatly improved outcomes for HER2-positive breast cancer, with most patients diagnosed at early stages now achieving a cure and experiencing fewer disease recurrences (von Minckwitz et al. 2017). However, in the metastatic setting, while approximately 16% of patients attain long-term disease control (Swain et al. 2015), the majority eventually develop resistance to anti-HER2 therapies, whether through primary resistance, where the disease progresses shortly after treatment initiation, or acquired resistance, where resistance emerges following an initial period of response.

The mechanisms underlying the resistance to HER2-targeted therapies are complex and multifaceted. HER2 mutations in the kinase domain can reduce the effectiveness of treatments by altering the receptor’s structure (Marín et al. 2023). Compensatory signaling pathways, such as upregulation of HER3 and IGF-1R or mutations in the PI3K/AKT pathway, allow cancer cells to bypass HER2 inhibition (Mishra et al. 2018; Nagata et al. 2004; Cizkova et al. 2013). Tumor cells can also evade the immune response, particularly antibody-dependent cellular cytotoxicity, by downregulating immune-recognition molecules or recruiting immunosuppressive cells (Loi et al. 2013). The tumor microenvironment, characterized by fibrosis or hypoxia, can also act as a physical barrier to drugs like Trastuzumab emtansine (T-DM1) and Trastuzumab deruxtecan (T-DXd) (Sonnenblick et al. 2020). Finally, some studies suggest that HER2 transcripts generated by alternative splicing code protein isoforms with enhanced dimerization and modification in their antibody (trastuzumab) binding domain, which are associated with this drug resistance (Scaltriti et al. 2007; Turpin et al. 2016). Thus, a deeper understanding of these resistance mechanisms remains a critical challenge in overcoming and managing breast cancer tumors expressing HER2.

Alternative splicing, a fundamental post-transcriptional process in eukaryotic gene expression, allows a single gene locus to produce multiple distinct mRNA transcripts, increasing protein diversity (Nilsen and Graveley 2010). In humans, over 95% of multi-exon (protein coding) genes are estimated to undergo alternative splicing, generating an average of seven mRNA isoforms and an expected similar number of proteins per gene (Pan et al. 2008). In the context of cancer, aberrant splicing has been implicated in various aspects of tumor biology, including drug resistance (Sveen et al. 2016; Marcelino Meliso et al. 2017).

In breast cancer, alternative splicing affects numerous genes involved in key cellular processes, including apoptosis, cell cycle regulation, and signal transduction (Yang et al. 2019). Specifically, HER2 alternative splicing has gained particular attention in the context of resistance to HER2-targeted therapies. HER2 splice variants have been identified in large scale (Veiga et al. 2022), but most of the studies have been focused on P95HER2 and delta16 HER2 (Δ16HER2) isoforms (Scaltriti et al. 2007; Turpin et al. 2016). The P95HER2 (also known as CTF611 or HER2-CTF; here and after, P95) is an incomplete isoform of the HER2 protein (molecular weight of approximately 95 kDa) lacking the extracellular protein domain and being constitutively active (Molina et al. 2001; Arribas et al. 2011). P95’s expression has been associated with poor prognosis and resistance to trastuzumab, as it lacks the antibody binding site (Scaltriti et al. 2007). The Δ16HER2, results from skipping HER2 exon 16, which encodes a small portion of the extracellular domain (Kwong and Hung 1998). The Δ16HER2 isoform assembles stable homodimers and is associated with increased transforming activity and metastatic potential (Turpin et al. 2016). Controversially, studies suggest that Δ16HER2 may contribute to trastuzumab resistance (Mitra et al. 2009), whereas others have found that it may enhance sensitivity to specific HER2-targeted therapies (Castagnoli et al. 2014). Thus, the complex interplay between HER2 splicing isoforms in the context of drug response and resistance requires a more extensive and in-depth investigation.

In this study, we systematically investigated HER2 alternative splicing isoforms in breast cancer and their potential role in mediating resistance to anti-HER2 therapies. Our findings expand in 3.5x the currently known repertoire of HER2 protein-coding isoforms, confirming their expression and translation in a large breast cancer cohort and characterizing the structural and functional properties of these HER2 splicing isoforms. In cell culture models treated with Trastuzumab, T-DM1, and T-DXd, we identified specific patterns of HER2 splicing isoform expression associated with drug resistance, which were shifted to express variants lacking key antibody-binding domains. Globally, our study provides new insights into the role of HER2 alternative splicing and highlights its importance in breast cancer and gene-target drug resistance.

## RESULTS

### Assessing the HER2 splicing isoform diversity in breast cancer

Globally, our study comprised five main steps (Figure 1A). First, we expanded the known repertoire of HER2 protein-coding splicing isoforms using long-read sequences from breast tumors. Second, we used computational models, classical and based on deep learning, to characterize the main features of the proteins encoded by these HER2 splicing isoforms, including their functional domains, structural elements, and cellular localization. Third, we analyzed the isoform’s expression profile and their translational evidence in a large set of primary breast tumors. Fourth, we evaluated HER2 isoform expression in breast cancer cell cultures, which were sensitive or resistant to trastuzumab, T-DM1, and T-DXd. Finally, we compared the expression patterns of HER2 isoforms before and after the emergence of resistance in cell lines exposed to T-DM1 treatment. To ensure reliable isoform expression data, we carefully selected and stratified samples based on technical considerations (*e.g.*, distinct library preparation strategies or low number of mapped reads), confounding effects (*e.g.*, excluding male samples), and biological information about the tumors (HR and HER2 status). We grouped the samples into hormone receptor-positive (HR+) and HR-negative (HR-) categories and further sub-grouped them based on their HER2 expression status: HER2-high (HER2+++ in immunohistochemistry, or FISH/ISH amplified), HER2-low (HER2+ or HER2++, without FISH/ISH amplification), and HER2-zero (no staining), as shown in Figure 1B. This stratification was necessary to ensure fair comparison among breast cancer subtypes, given the distinct evolution and prognosis expected for different patient groups.

**Figure 1.**
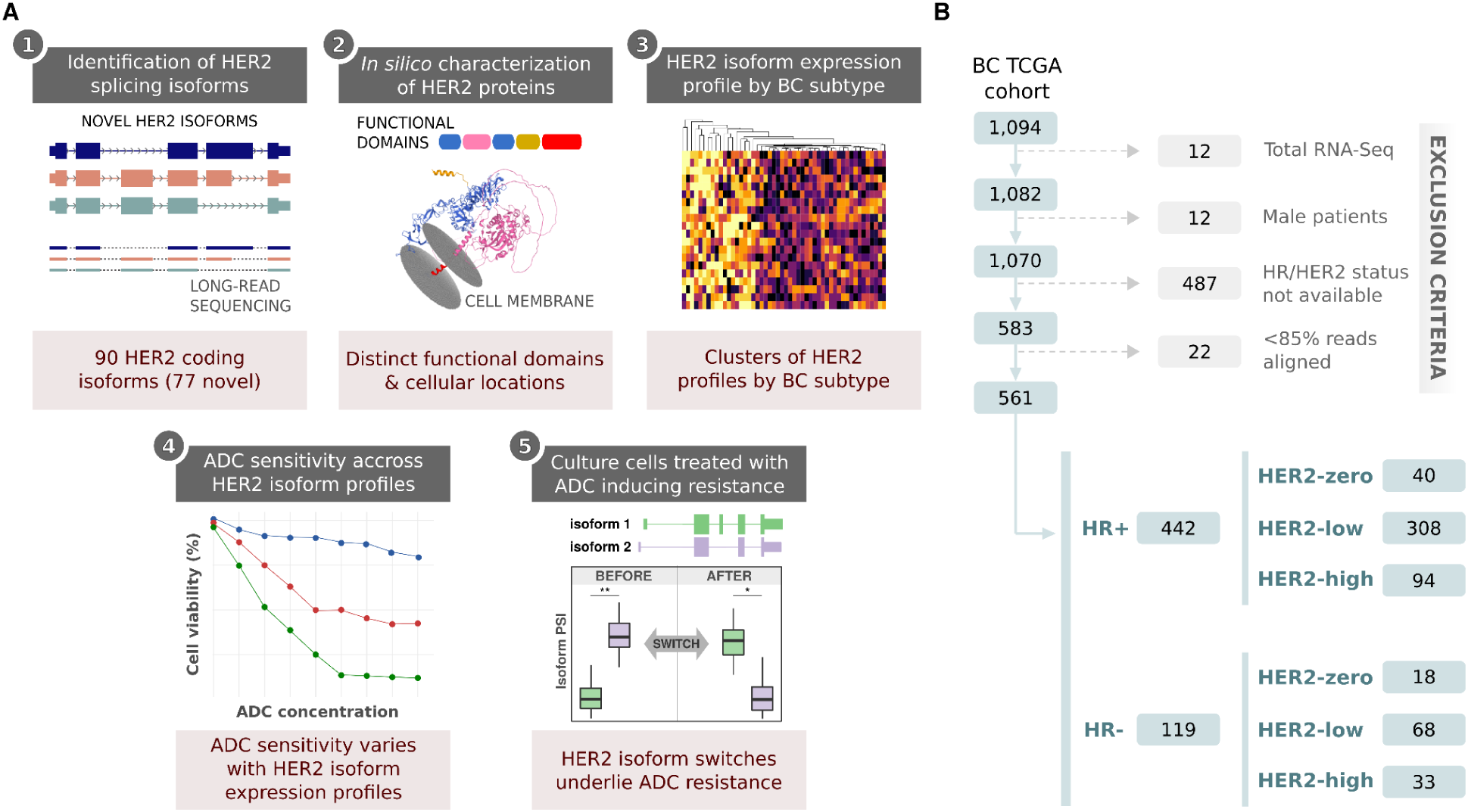
A comprehensive strategy for investigating HER2 isoform diversity in breast cancer. **A)** Five-step approach to characterize HER2 splicing isoforms in breast cancer patients and cell cultures: (1) Identification of HER2 splicing isoforms, (2) *In silico* characterization of HER2 protein variants, (3) HER2 isoform expression profiling by breast cancer subtype, (4) Analysis of antibody-drug conjugate (ADC) sensitivity across HER2 isoform profiles, and (5) Examination of HER2 isoform switches in ADC-induced resistance. **B)** RNA-sequenced samples selection and stratification based on their technical (*e.g.*, distinct library preparation strategies and low number of mapped reads) and biological characteristics (*e.g.*, male samples were excluded and tumors with same HR status and HER2 expression levels were grouped).

### Expanding the range of functional and structural variants of HER2 splicing isoforms

In the human reference transcriptome (GENCODE V36; (Frankish et al. 2019)), the HER2 (ERBB2) gene spans approximately 42.5 kilobases on chromosome 17 (chr17: 39,687,914 - 39,730,426; reference genome version GRCh38), comprising 27 canonical exons and 13 distinct protein-coding isoforms (Figure 2A). By using full-length mRNA transcripts (see Methods), we have significantly expanded the known repertoire of protein-coding HER2 splicing isoforms from 13 to 90 (Figure 2A). In terms of alternative splicing classes, these isoforms primarily result from exon skipping (SE, 40 isoforms), alternative 5’ splice sites (A5, 26 isoforms), alternative 3’ splice sites (A3, 15 isoforms), and other types (12 isoforms), Figure 2A. We observed alternative splicing events in specific HER2 exons, irrespective of the protein domains they encode. Twenty-three exons (85.1%; 23/27) of the canonical isoform showed evidence of alternative splicing (Figure 2A, colored exons; Supplementary Table 1).

**Figure 2.**
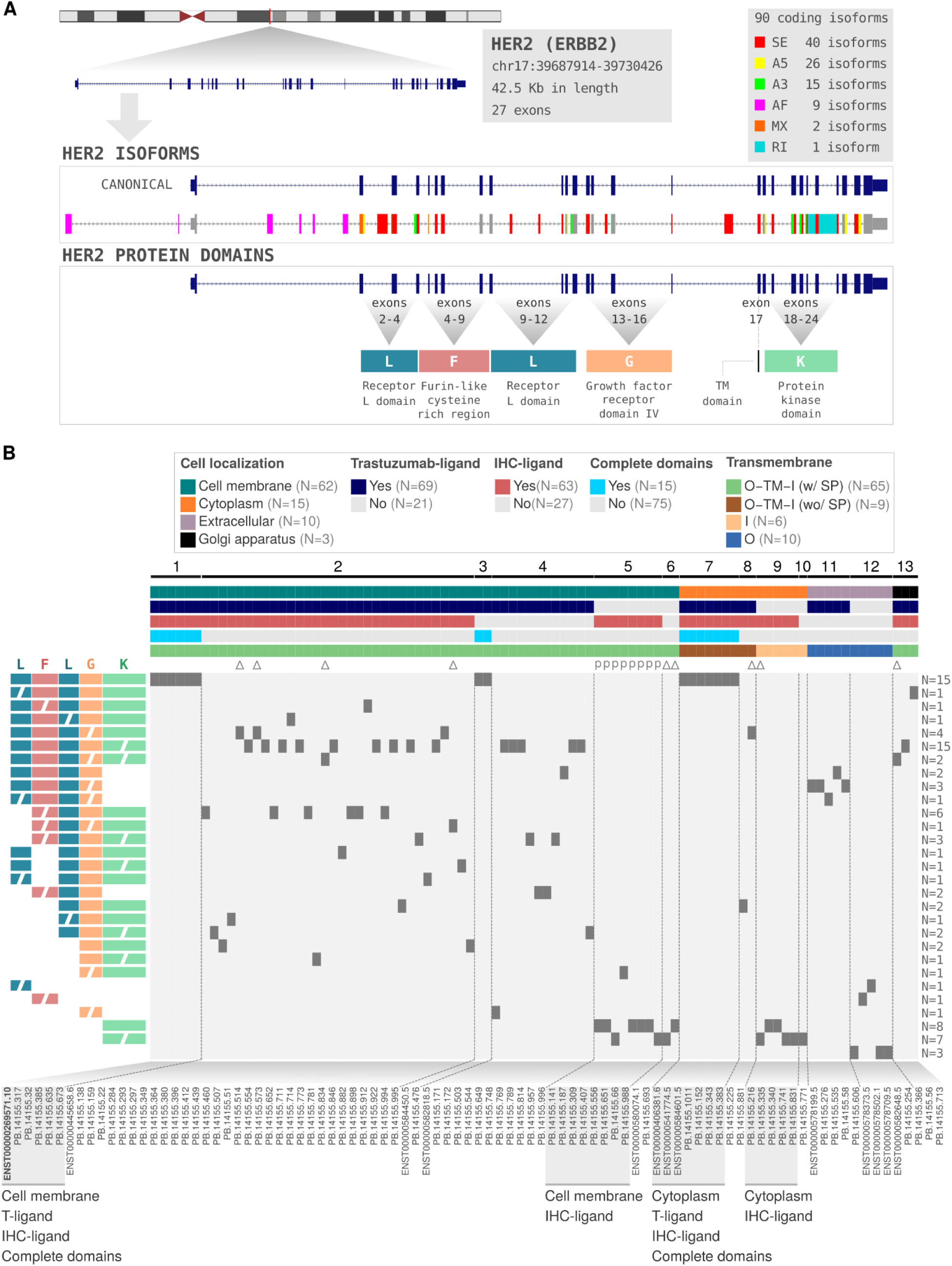
Comprehensive analysis of HER2 (ERBB2) isoforms and their characteristics. **A)** Genomic structure of the HER2 gene and its isoforms. The top panel shows the chromosomal location and exon structure of HER2. The middle panel displays the canonical HER2 isoform and alternative splicing events. The bottom panel illustrates the protein domains encoded by specific exons. **B)** Structural and functional properties of HER2 isoform-encoded proteins. The top bar represents the characteristics of isoform-encoded proteins (*e.g.*, cellular localizations, presence of trastuzumab and IHC binding region, transmembrane topology: O = outside, TM = transmembrane region, I = inside, SP = signal peptide) and the isoform-encoded protein groups, from 1 to 13 created based on their characteristics. The heatmap shows the presence (colored squares) or absence (white spaces) of specific protein domain configurations for each isoform-encoded protein. Protein domain configurations are on the left side and represented by colored letters (L: Receptor L domain, F: Furin-like domain, G: Growth factor receptor domain, K: Protein kinase domain). The presence of incomplete domains is represented by segmented labels. On the right side is the number of isoform-encoded proteins with each protein domain configuration. The canonical HER2 isoform (ENST00000269571) is highlighted in bold. P95 and delta16 isoforms are represented by "P" and "Δ", respectively. AS: alternative splicing; SE: skipping exon; AF: alternative first exon; A3: alternative 3’splice sites; A5: alternative 5’splice sites; MX: mutually exclusive exons; RI: retained intron.

To gain insights into their functional features, we categorized HER2 splicing isoforms into 13 distinct groups based on key characteristics of the proteins they encode: i) completeness of HER2 protein domains (Receptor L, Furin-like, Growth factor receptor and Protein kinase); ii) predicted cellular localization (cell membrane, cytoplasm, extracellular, or Golgi apparatus); iii) presence of the trastuzumab-binding domain; and iv) presence of the immunohistochemical (IHC) ligand domain (Figure 2B; Supplementary Table 2; see Methods for details). Each group represents isoforms sharing similar functional properties but displaying distinct combinations of these features. This classification system revealed substantial heterogeneity in HER2 isoform structural and functional characteristics.

Notably, the majority of isoforms (57.8%, 52/90; groups 1 to 4) retain cell membrane localization, but 21 (23.3%; groups 5, 6, 9, 10, and 12) lack the trastuzumab-binding site, including 10 isoforms also with cell membrane localization prediction (groups 5 and 6). Curiously, 35 (38.9%; groups 3, 4, 6, 10, 11, and 12) isoforms do not contain the binding domain of antibodies used for IHC staining, which may affect HER2 expression determination by IHC, a critical factor in breast tumor classification. Finally, only 15 isoforms (16.7%) retain all canonical protein domains, suggesting that most alternative isoforms may have some degree of altered functional properties (Figure 2B).

HER2 has two well-studied variants, P95 and Δ16HER2. Here, we took advantage of the full-length transcript analysis to examine beyond individual splicing events (*e.g.*, skipping exon), revealing complete transcripts that combine multiple alternative splicing events and may encode proteins different from those predicted by examining isolated events alone. Using this approach, we identified nine distinct isoforms that loses exon 16 (Δ16HER2 variants): only one reported in GENCODE (ENST00000580074.1) and eight novel isoforms discovered through our long-read sequencing strategy (Supplementary Figure 1A). These Δ16HER2 isoforms are distinct because they exhibited additional alternative splicing events, including alternative first exon usage, 5’ alternative splice site selection, skipping of exons 19 and 24, and, in one case, three additional alternative splicing events. For P95HER2 (P95), our analysis revealed eight splicing isoforms that encode P95-like proteins (Supplementary Figure 2A). These isoforms also demonstrated complex splicing patterns, including alternative first exon usage (1 isoform), exon skipping (1 isoform), combinations of alternative first exon with other splicing events (6 isoforms), and upstream small ORFs (see below).

Collectively, this comprehensive characterization reveals a complex and diverse landscape of HER2 splicing isoforms, extending even to well-studied variants like P95 and Δ16. It also provides a robust framework for generating new hypotheses about the functional roles of HER2 splicing isoforms and their potential impact on resistance to HER2-targeted therapies in breast cancer.

### Characterization of HER2 isoforms diversity at protein and mRNA levels

To further characterize the spectrum of HER2 splicing isoform diversity, we analyzed their features at both the mRNA (transcript) and protein dimensions (Figure 3; Supplementary Tables 2 and 3). Transcript length analysis revealed two significant sets of isoforms. First, isoforms with a primary peak length near the canonical HER2 transcript (ENST00000269571; 4,557 nt) and another minor peak around 2,600 nt (Figure 3A). Protein length distribution showed a significant peak corresponding to the canonical HER2 protein (1,255 amino acids) and a pronounced left-skewed distribution of shorter proteins (Figure 3B).

**Figure 3:**
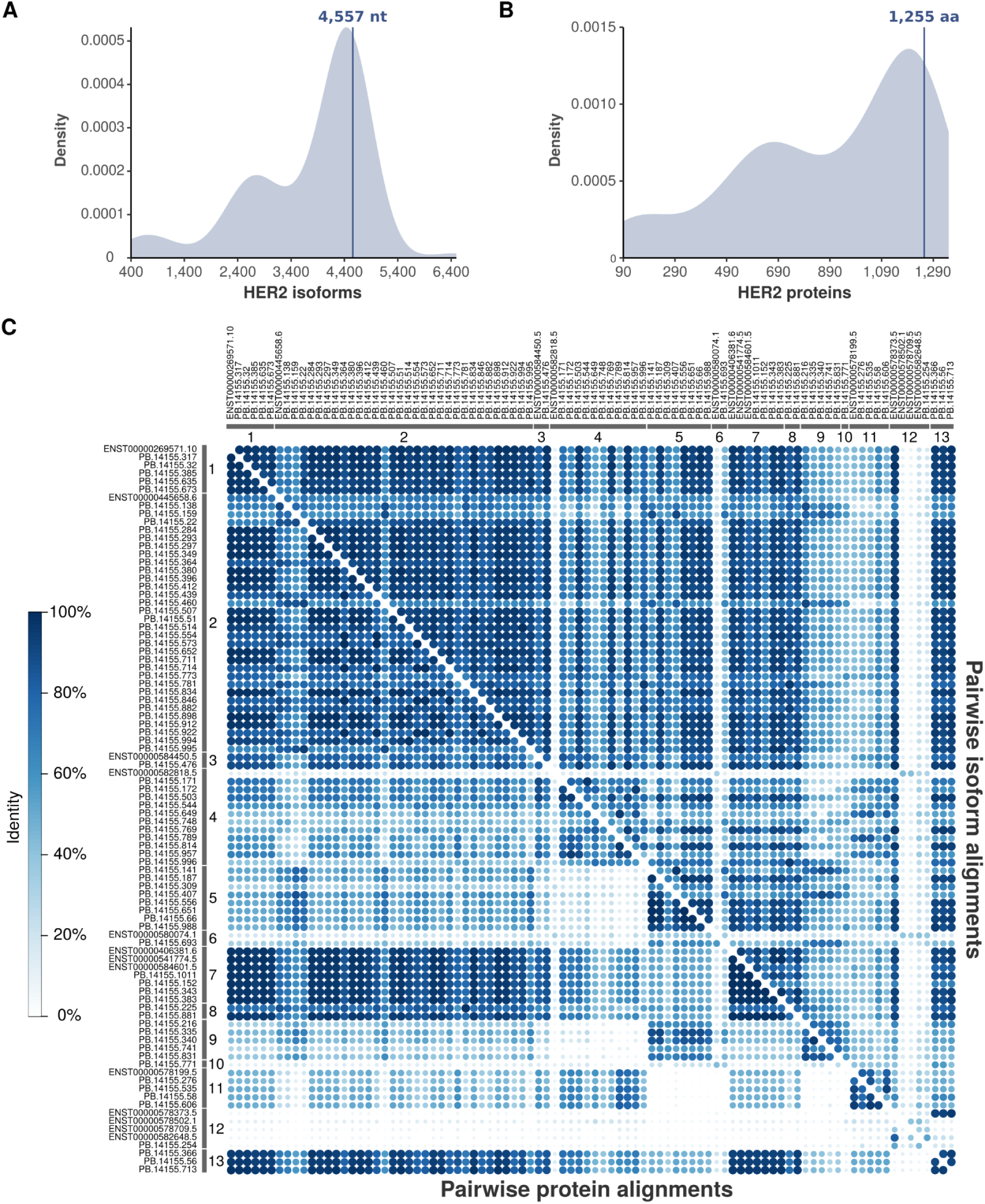
Distribution and pairwise alignment of HER2 isoforms. **A-B)** Density plots showing the distribution of HER2. **A)** transcript lengths (in nucleotides); and **B)** protein lengths (in amino acids). The vertical lines indicate the canonical HER2 transcript (4,557 nt) and protein (1,255 aa) lengths. **C)** Correlogram showing pairwise alignments of HER2 isoforms (upper diagonal) and proteins (lower diagonal). The color intensity represents the degree of sequence identity between isoforms/proteins, with darker blue indicating higher similarity. The diagonal represents self-alignments (100% identity). Isoforms are clustered based on similarity, and major groups are indicated by numbers 1-13.

To assess the degree of similarity among HER2 isoforms, we performed pairwise sequence alignments at both the transcript and protein levels (Figure 3C; Supplementary Table 4). The resulting correlogram revealed clusters of isoforms closely corresponding to the 13 functional groups defined in Figure 2B. Isoforms from groups 1, 2, 3, 7, 8, and 13 exhibited high similarity (>80%) at both nucleotide and amino acid levels, likely representing variants closely related to the canonical HER2 sequence (ENST0000026957, group 1). In contrast, most of isoforms from groups 4, 5, 6, 9, 10, 11 and 12 displayed lower similarity (<40% in most cases) to the canonical isoform and other groups.

We also evaluated nucleotides and amino acid similarities focused on the set of Δ16 and P95 HER2 splicing isoforms (Supplementary Figures 1B and 2B, respectively). While the eight P95 isoforms showed high similarities to each other at both transcript and protein levels, three of the nine Δ16 transcripts (33.3%, including the GENCODE-reported isoform ENST00000580074.1) exhibited low similarity to the other five isoforms. Thus, these results highlight the importance of analyzing HER2 diversity at transcript and protein levels.

### HER2 isoform expression patterns across breast cancer subtypes

After our comprehensive characterization of HER2 isoforms, we sought their expression across 561 breast cancer tumors from The Cancer Genome Atlas (Cancer Genome Atlas Network 2012) (Figure 1B). To have groups of clinically and biologically similar tumors and patients, we initially stratified tumors into six specific subgroups based on their hormone receptor (HR) status (HR+ or HR-) and HER2 expression levels (HER2-high, HER2-low, or HER2-zero) determined by IHC and/or FISH. The refined cohort was sub-classified in 442 samples HR+, including 40 with HER2-zero, 308 with HER2-low, and 94 samples with HER2-high status; 119 samples ER-, divided into 18 HER2-zero, 68 HER2-low, and 33 HER2-high samples (Figure 1B, Supplementary Table 5). Analysis of HER2 gene expression (pooled from all isoforms) using RNA-Seq data showed strong concordance with HER2 status determined by immunohistochemistry (Figure 4A and 4B). HER2-high tumors exhibited the highest expression levels, followed by progressively lower expression in HER2-low and HER2-zero tumors. This gradient of expression was observed in both HR+ (Figure 4A) and HR-(Figure 4B) tumors, with significant differences between all HER2 status categories (Mann-Whitney test, p-value < 0.05).

**Figure 4.**
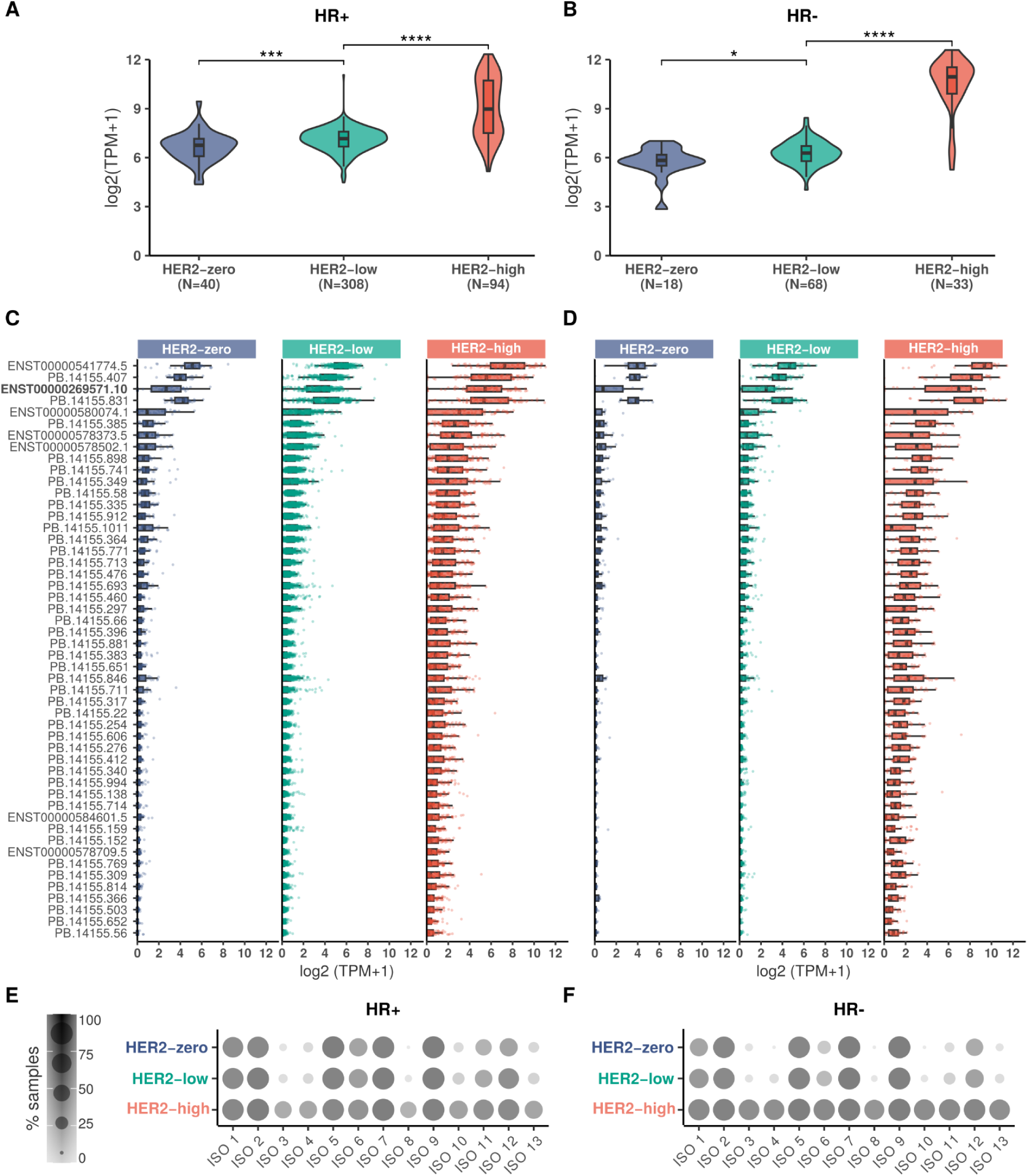
Expression profiles of HER2 gene and splicing isoforms in breast cancer samples classified by immunohistochemistry status of HER2 and hormone receptor. **A-B)** HER2 gene expression levels in **A)** HR+ and **B)** HR-breast cancer samples, stratified by HER2-high (red), HER2-low (green), and HER2-zero (blue) status. **C-D)** Expression profiles of the top 50 most expressed HER2 isoforms in **C)** HR+ and **D)** HR-breast cancer samples, stratified by HER2 status. The canonical HER2 isoform is highlighted in bold. Expression levels are shown in log2(TPM+1). Statistical significance: * p-value < 0.05, *** p-value < 0.001, **** p-value < 0.0001. **E-F)** Dot plots showing the percentage of **E)** HR+ and **F)** HR-breast cancer samples expressing each HER2 isoform group (ISO 1-13), stratified by HER2 status (zero, low, high).

Next, we analyzed the expression profiles of individual HER2 splicing isoforms across samples from distinct HR and HER2 statuses. Expression evidence was observed for all isoforms (Supplementary Figure 3). We then examined the expression patterns of HER2 isoforms in detail (Figures 4C (HR+) and 4D (HR-), the top 50 most expressed isoforms; Supplementary Figure 4 and Supplementary Table 6, all 90 isoforms). Overall, HER2-high patients presented the highest expression for all isoforms, followed by HER2-low. HER2-zero samples presented the lowest expression for all isoforms. Accordingly, all breast cancer subtypes presented the same top four most expressed isoforms, including the canonical isoform (ISO 1: ENST00000269571) (Figure 4C-D). Interestingly, however, the canonical isoform was not the highest expressed within all groups. In HR+ samples (Figure 4C), the canonical HER2 isoform exhibited a gradient of expression (median: HER2-high ∼6 TPM; HER2-low ∼ 4 TPM, HER2-zero ∼ 3 TPM). ENST00000541774.5 (ISO 7) and PB.14155.831 (ISO 9) showed consistently high expression across all HER2 categories (median > 6 TPM for all groups). PB.14155.385 (ISO 1) and ENST00000578373.5 (ISO 12) displayed a gradient similar to the canonical isoform, but with lower overall expression levels. In HR-samples (Figure 4D), while overall patterns of HER2 splicing isoforms were similar to HR+, some differences were highlighted. The median of expression of HER2-high versus HER2-low and -zero was more pronounced for multiple splicing isoforms, including the canonical. An increased expression variability was noted in the HER2-high group for specific isoforms (*e.g.*, ISO 1: PB.14155.385), and a subset of isoforms (*e.g.*, ISO 2: PB.14155.922, ISO7: PB.14155.343) showed detectable expression primarily in HER2-high HR-samples while remaining minimally expressed across all HR+ categories. Although some differences were observed, no distinct expression patterns were evident within patient groups.

Subsequently, we assessed the prevalence of (% of samples expressing) different HER2 isoform groups (ISO 1-13) across breast cancer subtypes, as shown in Figures 4E (HR+) and 4F (HR-). In HR+ samples, isoform groups ISO 1, 2, 5, 6, 7, 9, and 12 showed moderate to high prevalence across all HER2 categories (zero, low, and high). A similar overall pattern was observed in HR-samples, although with some differences. HER2-high tumors showed more uniform prevalence across all isoform groups than HER2-low and HER2-zero categories. ISO 1-7 were more expressed in HER2-high HR-samples. HER2-zero and HER2-low HR-samples showed more selective isoform expression, with ISO 1, 2, 5, and 6 being the most consistently expressed groups. In both HR+ and HR-, the HER2-high presented the highest number of isoforms expressed in the highest number of samples.

Given the established association between P95 expression and poor clinical outcomes, including drug resistance (Scaltriti et al., 2007; Arribas et al., 2011), we conducted a detailed investigation of isoforms encoding P95-like proteins. Specifically, we analyzed the presence of upstream open reading frames (uORFs), known regulatory elements that modulate protein expression through various mechanisms (Young and Wek 2016). Among the P95 HER2 isoforms identified, six (PB.14155.66, PB.14155.556, PB.14155.187, PB.14155.988, PB.14155.651, and PB.14155.309) contained both predicted ORFs and uORFs, while two (PB.14155.407 and PB.14155.1411) lacked uORFs (Supplementary Figure 5A). Expression analysis revealed significant differences between isoforms with and without uORFs across all HER2 status categories (Supplementary Figure 5B). Consistent with the typical repressive effect of uORFs on downstream protein expression (Lee et al. 2021), isoforms lacking uORFs showed significantly higher expression levels compared to those containing uORFs (p-value < 0.0001). This regulatory mechanism adds another layer of complexity to HER2 isoform expression and potentially influences their biological impact in breast cancer. Altogether, these results reveal a complex landscape of HER2 isoform expression across breast cancer subtypes, diverging from total HER2 levels.

### Mass spectrometry validation and structural prediction of HER2 isoform-derived proteins

Since expression evidence was observed for all identified HER2 splicing isoforms, we proceeded with their characterization at the protein level to strengthen their biological relevance and functionality. Mass spectrometry (MS) provided crucial validation by offering direct evidence of protein isoform expression. Using MS data from 76 breast tumors, we confirmed the presence of proteins from all isoform groups (Figure 5A, Supplementary Table 7). Consistent with RNA-seq data, HR+ tumors showed proportionally more proteins confirmed by MS than HR-tumors. Similarly, HER2-high tumors presented more validated proteins than HER2-low and HER2-zero tumors, with the latter showing the lowest level of validation (Figure 5A, bottom panel). While isoform groups 5, 6, 9, 10, 11, and 12 showed variable detection patterns, groups 1, 2, 3, 7, 8, and 13 were more frequently detected (Figure 5A, bottom panel).

**Figure 5.**
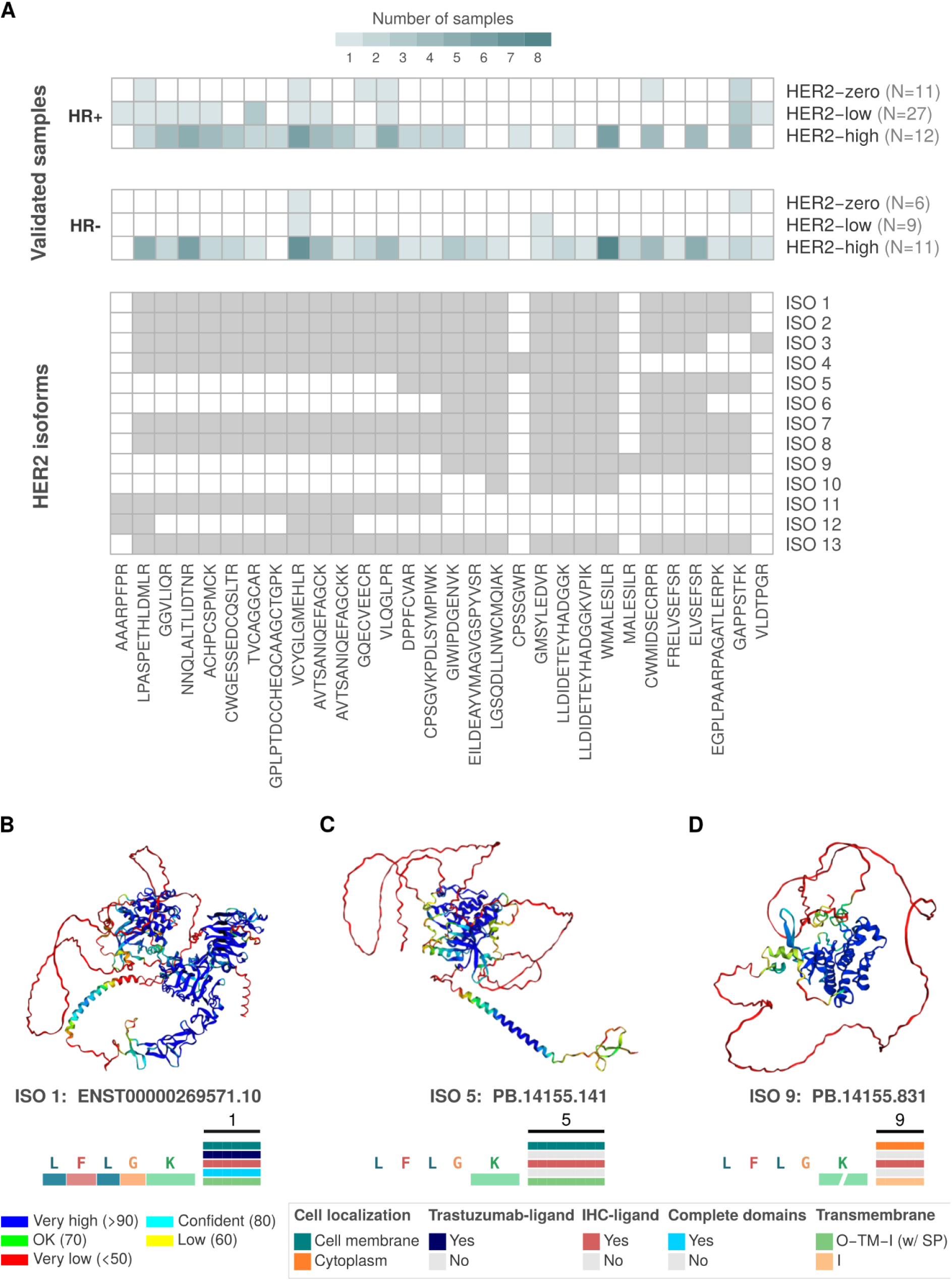
Mass spectrometry validation of HER2 isoform-derived proteins and their predicted 3D structures. **A)** The top panels display the number of samples with mass spectrometry (mass spec) confirmation of HER2 proteins, stratified by HER2 status (HER2-high, HER2-low, HER2-zero) and hormone receptor status (HR+ and HR-). Isoform-derived proteins are grouped from 1 to 13, and the validated peptides for each group are indicated in the lower panel. **B-D)** AlphaFold2-predicted protein structures for HER2 isoforms. **B)** shows the canonical isoform (ISO 1: ENST00000269571), with well-defined domain regions. **C)** displays ISO 5 (PB.14155.141), a variant from Group 5 with specific domain alterations affecting its cellular localization and trastuzumab binding potential. **D)** represents ISO 9 (PB.14155.831), an isoform with unique structural characteristics lacking complete domains, potentially affecting functional properties. Color codes in each structure represent pLDDT confidence scores for structural predictions and the corresponding HER2 protein domains, as the bottom legend indicates. Transmembrane: O = outside, TM = transmembrane region, I = inside, SP = signal peptide.

Next, we investigated the three-dimensional structure of protein isoforms, a fundamental step to understanding their function, interactions, and potential as therapeutic targets. For HER2 isoforms, structural information is particularly crucial as it can reveal how alternative splicing events may alter receptor conformation, ligand binding, dimerization, and downstream signaling capabilities. However, experimental determination of protein structures, especially for multiple isoforms, is time-consuming, expensive, and often challenging. The advent of AlphaFold has revolutionized the field of protein structure prediction (Jumper et al. 2021), allowing the investigation of protein isoforms at an unprecedented scale and speed. Using AlphaFold2, we found striking diversity in the predicted protein structures of HER2 isoforms which further corroborated our previous *in silico* predictions. The canonical HER2 isoform (ISO 1: ENST00000269571) displays the complete domain structure (transmembrane domain, full extracellular and intracellular domains, and juxtamembrane domain) with high confidence predictions across most regions (Figure 5B and Supplementary Figure 6). In contrast, ISO 5 (PB14155.141) shows significant structural alterations, particularly in the extracellular region (Figure 5C). Similarly, ISO 9 (PB14155.831) exhibits a dramatically altered structure, lacking several key domains (transmembrane and extracellular domains) and likely retaining only a partial kinase domain (Figure 5D).

Collectively, these results add a new layer of functionality to the HER2 isoforms by confirming their translation and presence in breast cancer samples and the structural predictions indicating their distinct functional properties.

### HER2 isoform clustering reveals novel subgroups within breast cancer patients

Next, we investigated the HER2 splicing isoform usage levels (Percent Spliced In - PSI values), which measure the relative abundance (expression) of a particular splicing isoform of a gene, across 561 breast tumors. First, we examined the internal variability in HER2 isoform PSI values per cancer subtype. This analysis revealed significant differences within variability in HER2 isoform expression patterns (Figure 6A). In HR+ patients, HER2-high samples showed significantly lower dissimilarity than HER2-low and HER2-zero samples (p-value < 0.0001; Mann Withney test), indicating more consistent isoform expression patterns in HER2-high tumors. HR-patients exhibited a similar trend, with HER2-high samples showing the lowest dissimilarity between HER2-low and zero (p-value < 0.05; Mann Withney test), Figure 6A.

**Figure 6.**
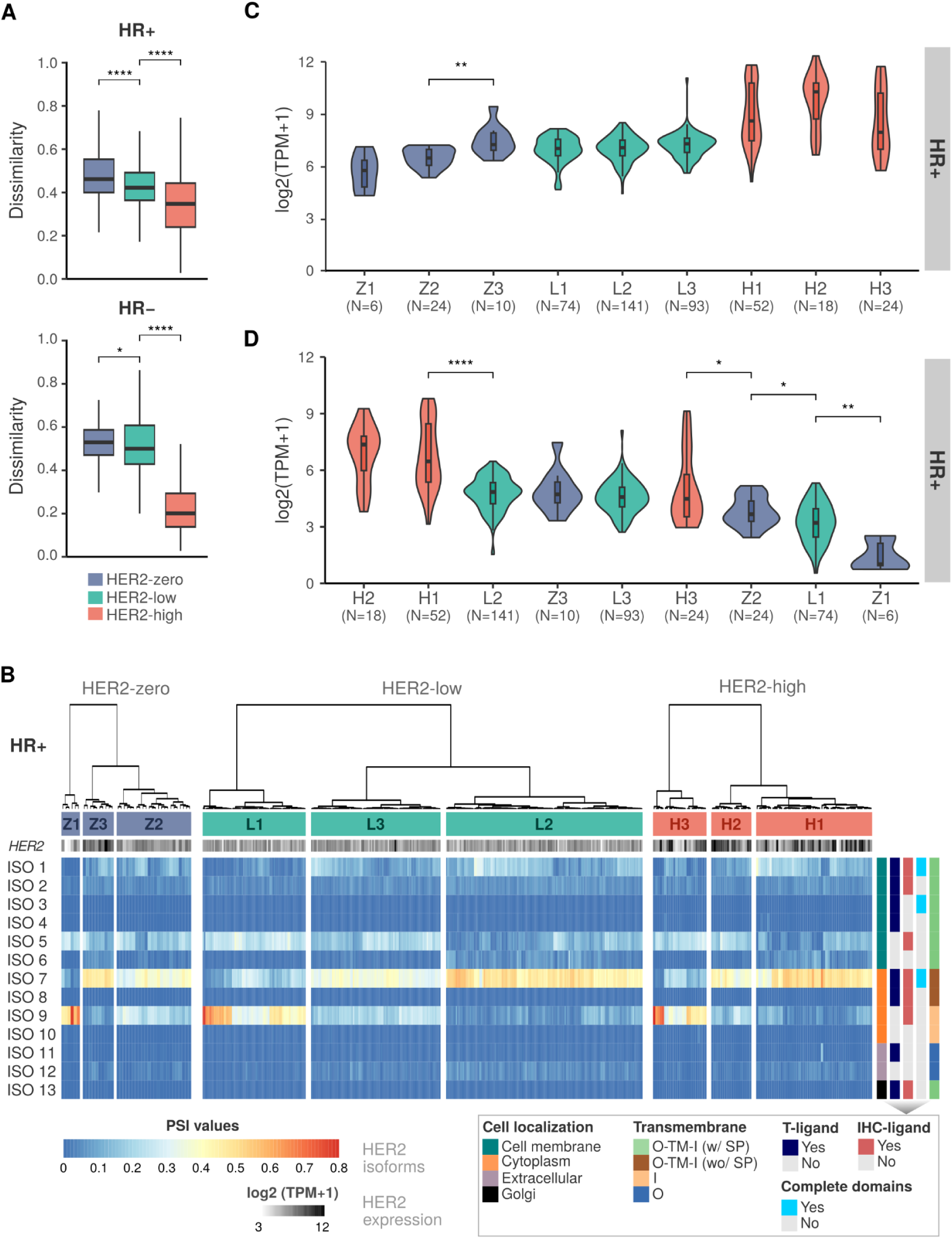
HER2 isoform expression patterns and clustering analysis in HR- and HR+ breast cancer patients stratified by HER2 status. **A)** Jaccard index comparing the dissimilarity of HER2 isoform expression between HER2-zero (blue), HER2-low (green), and HER2-high (red) breast cancer samples in hormone receptor-positive (HR+) and hormone receptor-negative (HR-) patients. Significant differences in dissimilarity are noted between HER2 status categories (* p-value < 0.05, **** p-value < 0.0001). **B)** Heatmap of HER2 isoform Percent Spliced In (PSI) levels in HR+ patients, clustered by expression similarity. Z1, Z2, Z3 for HER2-zero; L1, L2, L3 for HER2-low; H1, H2, H3 for HER2-high. Isoform features (cell localization, antibody binding sites, domain completeness and transmembrane topology: O = outside, TM = transmembrane region, I = inside, SP = signal peptide) are indicated on the right. **C)** HER2 gene expression levels among HR+ patient clusters. **D)** Expression levels of HER2 isoforms whose encoded proteins are located in the cell membrane and contain the trastuzumab-ligand (groups ISO 1-4) among HR+ patient clusters. Statistical significance: * p-value < 0.05, ** p-value < 0.01, **** p-value < 0.0001. Comparisons without statistical significance are not depicted in the figure.

Given this significant internal variability in isoform expression, we subsequently explored the possibility of identifying patient clusters exhibiting similar expression patterns within each HER2 subgroup. Accordingly, unsupervised hierarchical clustering of PSI values revealed distinct subgroups within clinically defined HER2 status categories (HR+ breast cancer samples: Figure 6B; HR-breast cancer samples: Supplementary Figure 7A; Supplementary Table 8). Notably, we found a complex landscape of HER2 isoform usage that extends beyond conventional HER2 expression levels. The clustering analysis identified three major clusters within each clinically defined tumor group for both HR+ and HR-patients: HER2-zero (clusters Z1-Z3), HER2-low (clusters L1-L3), and HER2-high (clusters H1-H3) (Figure 6B; Supplementary Figure 7A for HR-samples), revealing an intricate pattern of isoform expression that varied within tumor groups with the same HR/HER2 status, yet it was to some extent, recapitulated in clusters of distinct groups. Across all patients, isoforms from groups (ISO) 1, 5, 7, and 9 showed proportionally higher expression compared to all others, but with distinct expression patterns among them. Specifically, in HR+ clusters Z1, L1, and H3 (Figure 6B), as well as HR-clusters H3, L3, and Z2 (Supplementary Figure 7A), we observed high expression of ISO 9 and ISO 5. Since these isoforms lack the trastuzumab-binding domain, their predominance might predict poor response to HER2-targeted therapies. Conversely, HR+ clusters H1, L2, and Z3 (Figure 6B), along with HR-clusters H1, L2, and Z3 (Supplementary Figure 7A), showed highest expression of ISO 7 and ISO 1, which retain the trastuzumab-binding domain, suggesting potential favorable response to HER2-targeted therapies. Notably, clusters H2, L3, and Z2 (Figure 6B; H2, L1, and Z1 for HR-, Supplementary Figure 7A) exhibited a gradient of expression from ISO 7 to ISO 5, ISO 9, and ISO 1, suggesting more variable therapeutic responses.

Curiously, the HER2-low category exhibited the most heterogeneous isoform expression patterns. While the subgroup L3 closely resembled the HER2-high profile, the L1 showed isoform expression patterns more similar to HER2-zero tumors, enriched with expression of isoforms from group 9 (out of cell membrane and without trastuzumab ligand). This heterogeneity within the HER2-low category may explain the variable responses to ADC HER2-targeted therapies (T-DXd) observed in clinical studies of this patient population. To further characterize the HER2 expression patterns across the identified patient clusters, we analyzed both total HER2 gene expression and the expression of specific HER2 isoform groups. Total HER2 gene expression levels (*i.e.*, the sum of all isoforms), as expected, confirmed that HER2-high clusters (H1, H2, and H3) have an elevated HER2 expression (median 8-10 log2(TMP+1)) compared to the HER2-low (median 6.5-7.5 log2(TMP+1)) and HER2-zero (median 5.5-7 log2(TMP+1)) clusters (HR+, Figure 6C; HR-, Supplementary Figure 7B). Importantly, no statistically significant differences were found in total HER2 expression among clusters within each HER2 group, except for Z3 (HR+).

We next focused on the expression of HER2 isoforms encoding proteins localized to the cell membrane and containing the trastuzumab-binding domain (ISO 1-4 groups), given their potential clinical relevance for trastuzumab-based therapies (HR+, Figure 6D; HR-, Supplementary Figure 7C). The H2 and H1 clusters showed the highest expression of these clinically relevant isoforms. Remarkably, the H3 cluster (∼26% of HER2-high patients), despite being classified as HER2-high, exhibited lower expression of these isoforms (ISO 1-4), more closely resembling the levels seen in some HER2-low clusters. This result highlights the heterogeneity even within the HER2-high category and may suggest why HER2-high tumors have a low or no responsiveness to trastuzumab-based treatments. Notably, among the HER2-low, L2 (46% of HER2-low patients) and L3 (30% of HER2-low patients) show a moderated expression of ISO 1-4 isoforms, with levels between H1 and H3 clusters, which may support the fact that some HER2-low tumors are responsive to HER2 targeted ADCs, as T-DXd. In contrast, group L1 (∼24% of HER2-low patients) has low expression of ISO 1-4 and high expression of isoforms from groups 5 and 9 (which lack the trastuzumab binding domain), suggesting a potential group of lower/no response to T-DXd or other ADCs targeting HER2. The same pattern was observed for HR-samples (Supplementary Figure 7C). Finally, we also identified the most abundantly expressed isoforms from each isoform group (ISO 1-13) in both HR+ and HR-samples (Supplementary Figures 8 and 9, respectively). Overall, HR+ and HR-samples showed a similar expression pattern in terms of the most abundant expressed isoforms per group.

All together, this detailed isoform-level analysis reveals a complex picture of HER2 expression in breast cancers, identifying subgroups with distinct isoform utilization patterns that transcend traditional HER2 status classifications. These findings hold significant potential for refining patient stratification and improving response predictions to HER2-targeted therapies.

### HER2 isoform profiles concerning antibody-drug conjugate sensitivity

Next, we investigated the relationship between HER2 isoform expression patterns in breast cancer cell lines and their response to T-DM1 or T-DXd (Figure 7A; Supplementary Table 9). Among HR+/HER2+ cell lines, BT-474, EFM-192A, MDA-MB-361, and ZR-75-30 showed sensitivity to T-DM1, while UACC-812 was resistant. In HR-/HER2+ cells, AU565, HCC1954, MDA-MB-453, and SK-BR-3 demonstrated sensitivity to T-DM1, while JIMT-1 and UACC-893 were resistant. Interestingly, MDA-MB-453 and SK-BR-3 showed sensitivity to both T-DM1 and T-DXd, while BT-474 was resistant only to T-DXd. Notably, the BT-474 cell line contains a SLX4 mutation (c.1181G>C, p.R394T), previously identified as a potential mechanism of resistance to T-DXd, because SLX4 encodes a DNA repair protein that regulates structure-specific endonucleases and seems to play a role in resistance to TOP1 inhibition. Among HER2-cell lines, MCF7 and ZR-75-1 (HR+) and MDA-MB-231 (HR-) showed resistance to both ADCs, consistent with their low HER2 expression levels. These results indicate a complex relationship between HER2 (isoform) expression and ADC response. While most HER2+ cell lines expressing high levels of HER2 and both isoform groups (ISO 1-4 and ISO 5-13) showed sensitivity to ADCs, cell lines expressing lower levels of the isoforms from groups 1-4 (ISO 1-4; UACC-812, ZR-75-1, MDA-MB-231, MDA-MB-468, JIMT-1, BT-474), which have the trastuzumab-binding domains and are predicted to be located in cell membrane, are resistant to T-DM1 or T-DXd (Figure 7A).

**Figure 7.**
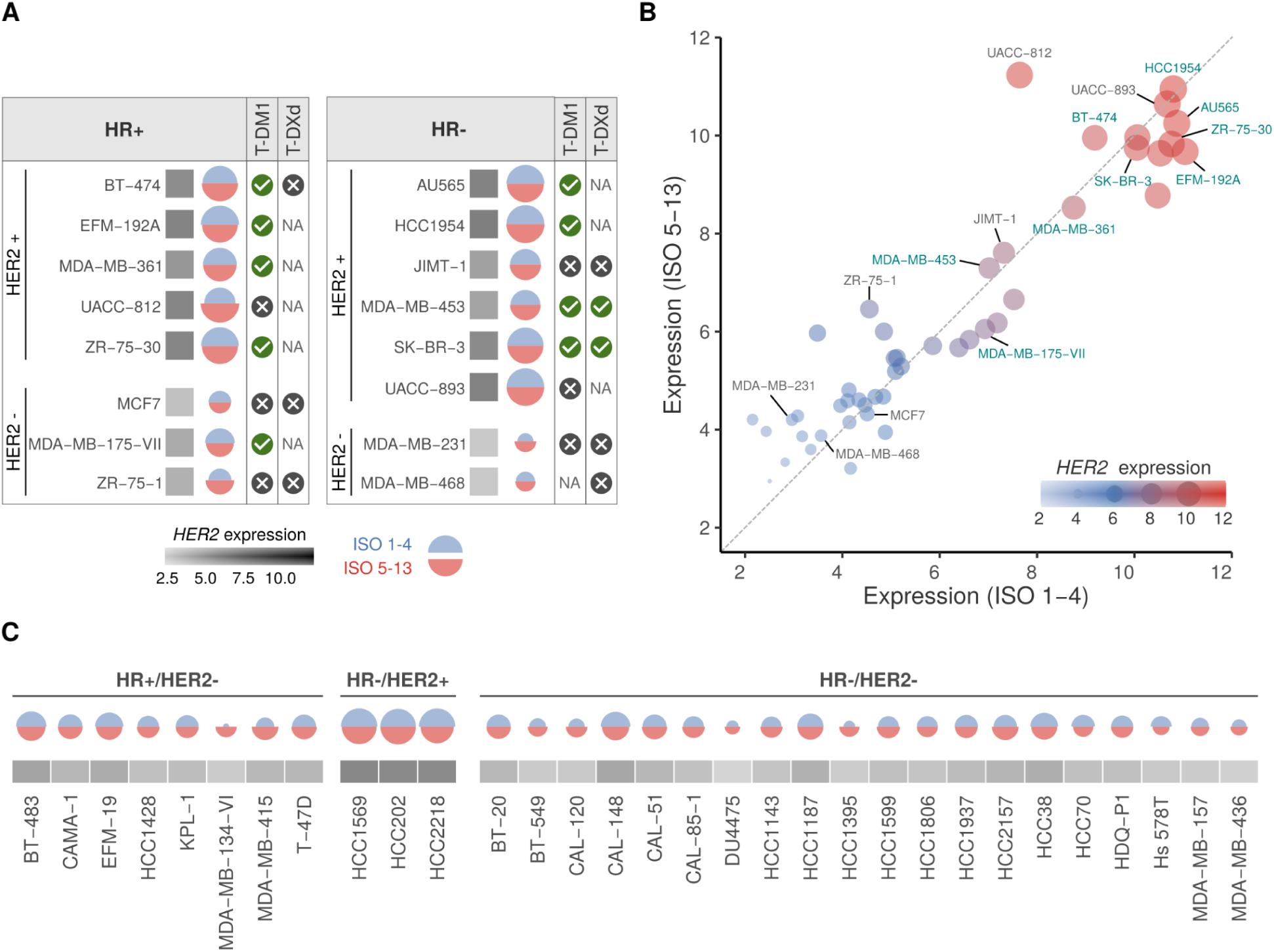
HER2 splicing isoform profiles in breast cancer cell lines and their response to T-DM1 and T-DXd. **A)** HER2 isoform expression in HR+ (left) and HR-(right) breast cancer cell lines treated with T-DXd or T-DM1. Cell lines responsive to treatment are marked with a "check symbol." Unresponsive cell lines are marked with a cross symbol. If drug treatment is not available: "NA". The square block’s color represents the total HER2 expression level (log2(TPM+1)). Blue semi-circular plots indicate the expression of isoforms with intact trastuzumab-binding domain (ISO 1-4). Red semi-circular plots half circus represent the expression of isoforms lacking trastuzumab-binding domain and/or cell membrane localization (ISO 5-13). **B)** Scatter plot showing the relationship between expression levels of ISO 1-4 and ISO 5-13 groups across breast cancer cell lines. Dot size and color intensity correspond to total HER2 expression level (log2(TPM+1)). Cell lines named in green indicate those responsive to antibody-drug conjugates (ADCs: T-DM1 or T-DXd); Cell lines named in gray indicate those ADC-resistant. **C)** HER2 isoform expression patterns across breast cancer cell lines (without ADC treatment) stratified by HR/HER2 status. Semi-circular plots represent expression levels of isoforms from groups 1-4 (blue) and 5-13 (red). The gray squares below indicate total HER2 expression levels.

Based on the previous results, we decided to better investigate the relationship between different HER2 isoform groups and ADC response. We evaluated the expression of ISO 5-13 (isoforms lacking trastuzumab-binding domain and/or located outside the cellular membrane) against ISO 1-4 (isoforms with intact trastuzumab-binding domain) across breast cancer cell lines (Figure 7B). Since the diagonal dashed line represents an equal expression of both isoform groups and the circle size indicates the levels of HER2 gene expression, most ADC-responsive cell lines (labeled in green, such as HCC1954, AU565, and BT-474) clustered in the high-expression region and maintain a balanced ratio between ISO 1-4 and ISO 5-13 expression. Interestingly, UACC-812, in spite of showing high overall HER2 expression, demonstrated resistance to ADCs and exhibited higher expression of ISO 5-13 relative to ISO 1-4 (positioning it above the diagonal). Similarly, JIMT-1, ZR-75-1, MDA-MB-231, and MDA-MB-468 showed resistance to T-DM1 and/or T-DXd and a higher expression of ISO 5-13 relative to ISO 1-4. On the other hand, MCF7 and UACC-893 showed ADC resistance regardless of their ISO 1-4 to ISO 5-13 ratio. Notably, we have several different cell lines under and above the diagonal (each circle represents a cell line). Still, for most of them, we have no ADC treatment available (circles without names). To gain further insight into the expression profile of HER2 and its isoforms in these additional cell lines, but without ADC treatment information, we created Figure 7C. HR+/HER2− cell lines showed consistently low to moderate expression of both isoform groups, with relatively balanced ratios between ISO 1–4 and ISO 5–13. HR−/HER2+ cell lines exhibited the highest total levels of HER2 expression and maintained substantial expression of both isoform groups. The largest group, HR−/HER2− cell lines, demonstrated consistently low expression of both isoform groups, albeit with some variability in the relative ratios of ISO 1–4 and ISO 5–13.

Altogether, these results indicated a balanced expression of ISO 1-4 and ISO 5-13 in most cell lines, suggesting that this balance may be necessary for normal HER2 function and that its disruption, as observed in some drug-resistant lines (*e.g.*, shifted to ISO 5-13), may contribute to altered cellular responses to HER2-targeted therapies.

### Dynamic changes in HER2 isoform expression associated with acquired resistance to trastuzumab and T-DM1

Finally, to illuminate the molecular mechanisms underlying acquired resistance to HER2-targeted therapies, we investigated the HER2 gene and isoform expression profiles in breast cancer cell lines before and after developing resistance to trastuzumab and T-DM1 (Figure 8, Supplementary Table 10).

**Figure 8.**
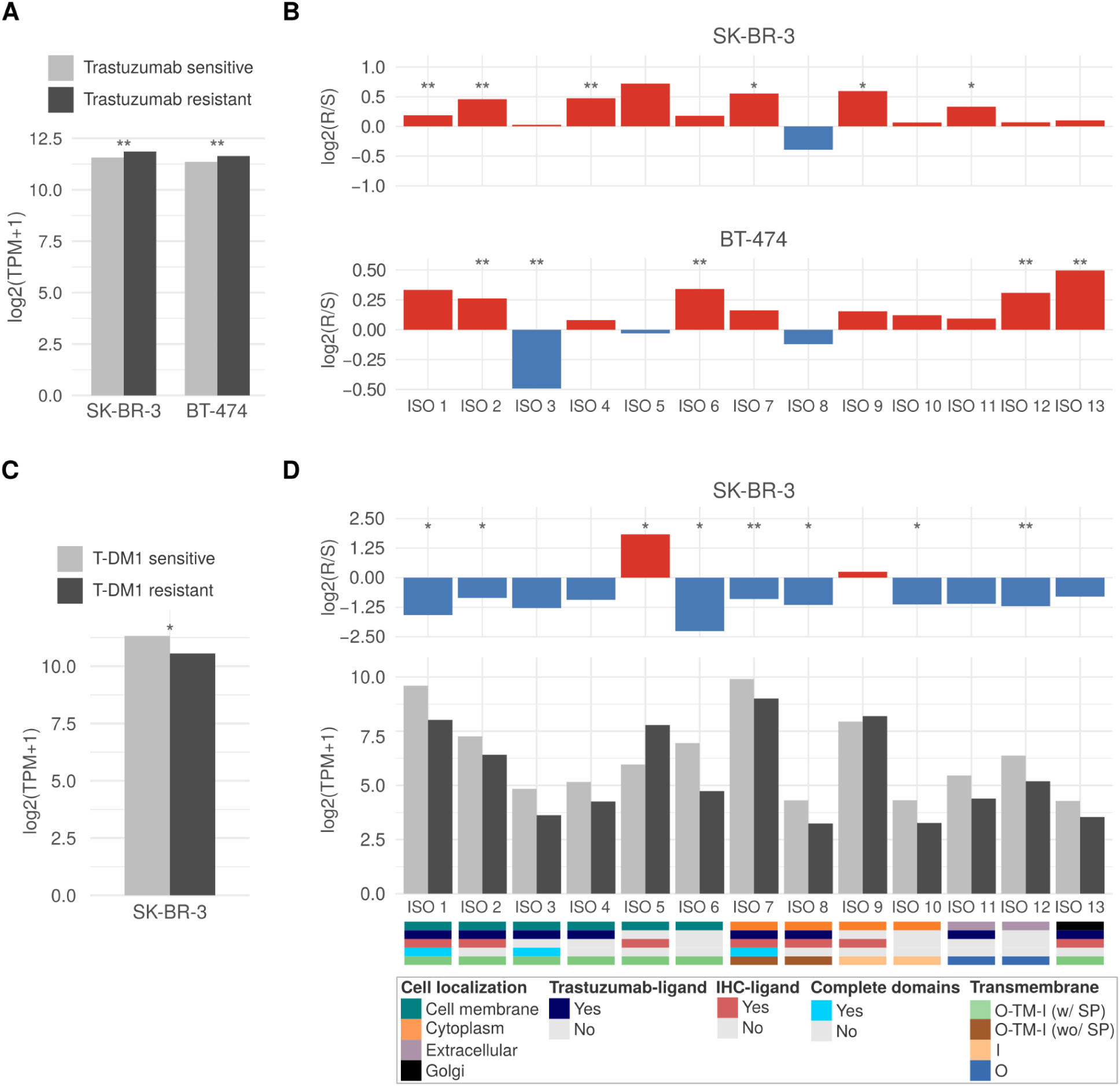
HER2 gene and isoform expression profiles in breast cancer cell lines before and after acquiring resistance to HER2-targeted therapies. **A)** Overall HER2 expression in trastuzumab-sensitive and -resistant SK-BR-3 and BT-474 cells. **B)** Log2 fold change of HER2 isoform expression in trastuzumab-resistant vs. sensitive SK-BR-3 and BT-474 cells. **C)** Overall HER2 expression in T-DM1-sensitive and -resistant SK-BR-3 cells. **D)** HER2 isoform expression levels and characteristics in T-DM1-sensitive and -resistant SK-BR-3 cells, with log2 fold change shown above. Isoforms are categorized based on their cell localization, trastuzumab-binding ligand presence (T-ligand), structural completeness and transmembrane topology: O = outside, TM = transmembrane region, I = inside, SP = signal peptide, as indicated by the bottom color legend. Statistical significance: * p-value < 0.05, ** p-value < 0.01. Comparisons without statistical significance are not depicted in the figure.

We first examined the overall HER2 expression in SK-BR-3 and BT-474 cell lines in sensitive and resistant states to trastuzumab (Figure 8A). Interestingly, both cell lines in the two states maintained significantly high levels of HER2 expression (Figure 8A), indicating that resistance to trastuzumab is not primarily mediated by a global downregulation of HER2 expression (Vernieri et al. 2019). In fact, since trastuzumab targets HER2 function, we hypothesize that its upregulation may serve as a compensatory mechanism to offset its own inhibition by treatment, thus sustaining downstream signaling pathways that promote rapid tumor cell growth and proliferation.

Next, to gain deeper insights into potential resistance mechanisms, we analyzed the HER2 isoform expression profiles and fold change of each isoform group in trastuzumab-resistant versus sensitive cells for both SK-BR-3 and BT-474 lines (Figure 8B; Supplementary Figure 10). In SK-BR-3 cells, we observed significant upregulation of multiple isoform sets. In contrast, isoforms from group 8 showed downregulation (log2 fold change ∼ -0.4) in resistant conditions. Overall, BT-474 cells also exhibited upregulation of sets of isoforms, except for isoforms from groups 3, 5, and 8.

Next, we investigated the impact of acquired resistance to T-DM1 on HER2 expression in SK-BR-3 and BT-474 cells (Figure 8C; Supplementary Figure 11A). Unlike trastuzumab-resistant cells (Figure 8A), T-DM1-resistant SK-BR-3 and BT-474 cells showed a significant decrease in overall HER2 expression compared to sensitive cells, from 11.3 to 10.6 and 10.9 to 10.1 log2(TPM+1), respectively (Figure 8C; Supplementary Figure 11).

Lastly, we performed a detailed analysis of HER2 isoform expression levels in T-DM1-resistant and -sensitive SK-BR-3 and BT-474 cells. We observed complex changes in the isoform landscape for SK-BR-3 (Figure 8D) and BT-474 (Supplementary Figure 11B). First, the fold change (T-DM1 resistant/sensitive) analysis showed that most splicing isoforms are significantly downregulated, including isoforms from sets 1 to 4 (isoforms with intact domains; Figure 8D). Remarkably, splicing isoforms from sets 5 and 9 are upregulated in T-DM1-resistant cells (Figure 8D). In BT-474, we observed a significant downregulation of all splicing isoforms (Supplementary Figure 11B). Altogether, these findings suggest that SK-BR-3 has adapted to treatment pressures by altering the balance of HER2 isoforms to downregulate the drug target isoforms (groups ISO 1 to 4) and upregulating the pro-survival signaling (isoforms from groups ISO 5 and 9, which lack the trastuzumab binding site and seems to retain the signaling capabilities through the tyrosine kinase domain), a putative mechanism to evade this ADC’s effects.

## DISCUSSION

In this investigation, we uncover a complex landscape of HER2 splicing isoform diversity in breast cancer that goes well beyond the conventional understanding of HER2 biology (Arteaga and Engelman 2014) and its role in targeted therapies (Modi et al. 2020; Tarantino et al. 2020). The full characterization of the set of 90 HER2 coding isoforms, including 77 novel variants, significantly expands our knowledge of HER2 expression and variations and sheds light on the role of HER2 splicing isoforms in antibody-conjugated targeted therapy resistance.

First, our strategy emphasizes the importance of identifying alternative splicing isoforms through the use of full-length transcripts obtained via long-read sequencing. This approach has allowed us to achieve a comprehensive characterization at the isoform level, rather than solely focusing on the splicing events themselves. This reveals the composition of all exons and the open reading frame (and subsequent protein) encoded by each isoform. The breadth of our strategy becomes clear when we look at the two most studied HER2 isoforms, Delta16 and P95: i) identified 8 distinct isoforms with different splicing events that encode proteins lacking the extracellular domain and have an approximate molecular weight of 95 kDa; ii) for Delta16, we found 9 isoforms, all containing the exon 16 skipping (Delta16’s hallmark) and exhibiting other alternative splicing events. Understanding not just the event (*e.g.*, skipping exon), but the full set of isoforms that contains such events and others certainly gives us a more complete understanding of the importance and functionality of each alternative splicing isoform.

The structural and functional diversity revealed in the HER2 isoforms, including alterations in the HER2 protein domains and cell localization, and the presence or lack of the trastuzumab binding sites, provides new insights into the heterogeneity of response in targeted therapy using antibodies and ADCs. These splicing isoform diversity profiles may explain, in part, the complex mechanisms of resistance to HER2-targeted therapies observed in clinical practice (Nahta and Esteva 2006; Luque-Cabal et al. 2016).

Curiously, we observed variability in the expression of HER2 splicing isoforms across the intrinsic subtypes of breast cancer, with the HER2-high group displaying the most uniform expression profile. Remarkably, this is the same patient group that shows the most consistent and profound response to anti-HER2 therapies, whether in early-stage or metastatic disease (Gianni Luca et al. 2012; Baselga Jose. et al 2012)). This correlation suggests that the homogeneity in HER2 isoform expression may play a role in influencing therapeutic response.

Identifying HER2 splicing isoforms lacking the antibody (drug) binding domains but retaining the signaling capabilities (tyrosine kinase domain), may explain a potential resistance mechanism to antibody-based therapies like trastuzumab and ADCs. This aligns with previous studies on P95, which lacks the trastuzumab binding site and has been associated with poor prognosis and resistance in antibody-based therapy treatment (Scaltriti et al. 2007). Therefore, the expanded repertoire of HER2 isoforms presented here suggests that alternative splicing may be a more prevalent and leading mechanism used by cancer cells in acquiring resistance and progression, especially in gene-targeted therapies.

Interestingly, the dynamic changes in HER2 isoform expression observed in cell lines acquiring resistance to trastuzumab or T-DM1 highlight the adaptive nature of cancer cells and may resemble the mechanism of therapy evasion in breast cancer patients. Specifically, our finding indicates that the SK-BR3 cancer cell line has adapted to treatment pressures by altering the balance of HER2 isoforms, downregulating those containing the drug target isoforms (isoforms in sets 1 to 4) and upregulating the HER2 splicing isoforms (sets 5 and 9) lacking the antibody (trastuzumab) binding site domains, which could be a key mechanism of acquired resistance in tumors under ADCs treatment. Broadly, these findings open new avenues for understanding and potentially mitigating therapy resistance through various strategies, including the development of HER2 isoform-specific inhibitors, combination approaches targeting multiple isoform-encoded proteins, and the use of tyrosine kinase inhibitors such as lapatinib that target the kinase domain.

Furthermore, our observations indicate that among tumor subtypes (HER2-high, -low, and -zero), there are distinct subgroups of tumors expressing different splicing isoforms, some of which encode protein isoforms lacking the binding domain for antibodies used in immunohistochemistry. This may explain why a percentage of HER2-low or even HER2-zero patients (as determined by immunohistochemistry) respond to treatment. It is reasonable to hypothesize that specific isoform compositions may create a false classification of HER2 status and vulnerabilities to ADC treatment, even in contexts with low HER2 expression (by immunohistochemistry) (Tarantino et al. 2020).

While our study provides comprehensive insights into HER2 isoform diversity and its implications for targeted therapy, several limitations should be acknowledged. First, our cell line-based resistance models, while informative, may not fully recapitulate the complexity of resistance mechanisms in breast cancer patients, where tumor heterogeneity and microenvironment factors play crucial and yet incompletely understood roles <u>(Vander Velde et al. 2020; Roma-Rodrigues et al.</u> <u>2017)</u>. Second, although we validated the existence of HER2 isoforms through RNA-seq expression in a large patient cohort and mass spectrometry confirmation, functional validation of individual isoforms’ biological roles and their specific contributions to drug resistance mechanisms requires further investigation. Third, while long-read sequencing enabled comprehensive isoform identification, technical limitations in detecting low-abundance transcripts might have led to underestimation of rare isoforms <u>(Uapinyoying et al. 2020)</u>. Fourth, our study focused primarily on the role of HER2 isoforms in antibody-based therapy resistance, and their potential impact on other treatment modalities, such as tyrosine kinase inhibitors, needs to be fully explored. Finally, while our findings suggest the importance of isoform-specific testing in clinical settings, the development and validation of practical diagnostic tools for HER2 splicing isoform profiling will require additional technical and clinical validation studies <u>(Wang and Aifantis 2020)</u>.

In conclusion, our comprehensive investigation into the diversity of HER2 isoforms uncovers a complex landscape that may have significant implications for breast cancer biology and treatment approaches utilizing antibody-drug conjugates (ADCs). Our findings indicate that integrating HER2 isoform profiling into clinical practice - despite its current limited implementation in many centers - may greatly improve patient stratification and treatment selection, potentially leading to more effective targeted therapies. This research establishes a solid foundation for a more refined approach to HER2-positive breast cancer. We propose that optimal ADC treatment strategies should be tailored not only to HER2 expression levels but also to the specific isoform profiles present in each tumor.

## METHODS

### Public RNA sequencing data

We obtained unprocessed RNA sequencing (RNA-Seq) data from 561 primary tumors sourced from female breast cancer patients, publicly accessible via The Cancer Genome Atlas (TCGA) repository (https://portal.gdc.cancer.gov). Additionally, we obtained clinical data detailing immunohistochemical (IHC) staining results for HER2, estrogen receptor (ER), and progesterone receptor (PR), as well as fluorescence in situ hybridization (FISH) data for HER2. In addition, RNA-Seq data from 50 breast cancer cell lines were acquired from the Cancer Cell Line Encyclopedia (CCLE, https://sites.broadinstitute.org/ccle/), and data on cell line sensitivity to T-DXd and T-DM1 were obtained from previous studies (Supplementary Table 9). RNA-Seq data from trastuzumab-sensitive and -resistant SK-BR-3 and BT-474 cell lines were obtained from (Duan et al. 2024) and (Mukund et al. 2024), respectively. Finally, RNA-Seq data from T-DM1-sensitive and -resistant SK-BR-3 and BT-474 cell lines were obtained from (Gedik et al. 2024)

### Quantification of isoform and gene expression levels

Using the Kallisto tool (version 0.48.0; default parameters with option --bootstrap-samples 100) (N. L. Bray et al. 2016), we pseudo-aligned the RNA sequencing reads from all patients and cell lines to an expanded version of the human reference transcriptome, comprising 13 known protein-coding HER2 isoforms from GENCODE (version 36; https://www.gencodegenes.org/human/) in addition to a set of 77 novel protein-coding HER2 isoforms identified through long-read RNA-Seq of breast cancer samples from (Veiga et al. 2022). Next, isoform expression levels normalized in transcripts per million (TPM) were submitted to SUPPA2 (version 2.3; default parameters) (Trincado et al. 2018), which quantifies percent spliced-in (PSI) values, indicating the proportion of expression that each isoform of a gene corresponds to. Gene-level expression profiles were also obtained using the tx_import R package (version 1.26.1) (Soneson, Love, and Robinson 2015).

### Characterization of HER2 splicing isoforms

To characterize the HER2 isoforms in terms of alternative splicing local events, coding potential, functional domains, transmembrane topology, and subcellular localization, we used several strategies. First, coding sequences (ORFs) from 77 novel HER2 isoforms previously determined (Veiga et al. 2022) using Transdecoder (Haas et al. 2013) were extracted, and coding sequences from 13 known HER2 isoforms were directly retrieved from GENCODE (version 36). Local alternative splicing events were identified using the “generateEvents” function in SUPPA2 (Trincado et al. 2018) with default parameters; for retained intron (RI) events, additional parameters --boundary V and --threshold 10 were applied. Multiple skipped exon (SE) events, which are not reported by SUPPA2, were manually extracted. Protein domains from all HER2 coding sequences were predicted using the hmmsearch tool from HMMER (version 3.3.1; default parameters) (Potter et al. 2018) based on the Pfam database (Mistry et al. 2021). Predictions of transmembrane topology were performed using the DeepTMHMM web tool (default parameters) (Hallgren et al. 2022), which uses a deep learning algorithm to predict the topology of alpha-helical and beta barrels. Protein subcellular localizations were determined based on the DeepLoc 2.0 tool (default parameters) (Ødum et al. 2024). To evaluate the presence of the immunohistochemical (IHC)-binding region in HER2 isoforms, we considered 3 IHC antibodies: PATHWAY HER2, Herceptest and Oracle HER (Cho et al. 2003). Besides, similarities among HER2 proteins were evaluated through pairwise protein alignments using the needle global aligner (https://www.ebi.ac.uk/jdispatcher/psa/emboss_needle).

### Validation of HER2 isoforms at the protein level

Validation of HER2 isoforms at the protein level was performed in MS/MS data from 76 breast cancer patients from the CPTAC TCGA (study ID: PDC000173) using the PepQuery tool (v2.0.2) (Wen and Zhang 2023) with default parameters. HER2 peptides from expressed splicing isoforms were queried in MS/MS data to find supporting peptide spectrum matches (PSMs). Only PSMs that passed all PepQuery filtering steps with FDR < 0.05 were considered confident. All proteins from the human database GENCODE release 36 and the predicted HER2 protein sequences from long-read data were used as references in the validation.

### Prediction of 3D protein structures of HER2 isoforms

The prediction of the HER2 isoforms’ 3D protein structures was made with AlphaFold2 (Jumper et al. 2021) through the free and publicly available Google collaborator ColabFold (v1.5.5) platform (Mirdita et al. 2022). We opted to run the predictions this way due to its speed by combining it with a fast homology search with MMseqs2 (Steinegger and Söding 2017) and HHsearch (Steinegger et al. 2019), and the usage of the highly accurate PDB100 (Varadi et al. 2024) as its database. All analyses were run in a “High-RAM (system: 51Gb; GPU: 15Gb) T4 GPU” machine with Python 3 and more than 200Gb of disk space.

All parameters were left default, except “num_recycles = 24” because membrane proteins require a higher number of recycles for better results. Several outputs are made available, including not only the predicted protein structure itself, but also alignments for reference, PDB files per ranked model for editions, and other plots to support the results. The quality of the predictions was assessed by analyzing two metrics: (i) the MSA (multiple sequence alignment) coverage outputs, where at least 30, ideally 100 sequences per position are ideal for better performance; (ii) the pLDDT scores, both for each aminoacid and also for the entire structure, where higher scores (out of 100) - ideally above 70% (“ok”), especially above 80% (“confident”) - mean more confidence and, as a consequence, better models (Supplementary Figure 6). The best model (rank 1, among five runs in total), *i.e.*, the one with the highest pLDDT score, was always chosen.

## Data Availability

All data produced in the present study are available in the manuscript or can be requested to the authors

## Acknowledgments

This work was supported by grant 2018/15579–8, São Paulo Research Foundation (FAPESP) to PAFG; grants 2020/14158-9 (to FFS), São Paulo Research Foundation (FAPESP). GDAG. was supported by a fellowship from the Young Scientist program, Hospital Sírio-Libanês. It was also partially supported by funds from CNPq (PAFG, AAC), Serrapilheira Foundation (PAFG and AB), and Hospital Sírio-Libanês to PAFG and AAC.

## Author contributions

G.D.A.G., CH dos A, and P.A.F.G. developed the concepts in this study. P.A.F.G. supervised the study. Analyses were performed by G.D.A.G. and CH dos A. The manuscript was written by G.D.A.G., CH dos A, and P.A.F.G. with contributions and revisions from all authors.

